# Policy gaps regarding social homecare in the context of end-of-life; a policy document analysis

**DOI:** 10.1101/2025.07.14.25331508

**Authors:** Helene Louise Elliott-Button, Kasonde Mwaba, Zana Bayley, Joan Bothma, Cat Forward, Jamilla Hussain, Justine Krygier, Mark Pearson, Paul Taylor, Caroline White, Jane Wray, Liz Walker, Miriam Johnson

## Abstract

**Background:** Social homecare workers (personal aides/assistants) are crucial for people wishing to receive end-of-life care at home.

**Aim:** To determine current UK social care policy priorities and gaps regarding end-of-life care provision by homecare workers, including support and training for this workforce.

**Design:** Policy document review and content analysis; eligible documents were current UK policy documents informing homecare worker practice/service delivery, identified by team expertise, networks, study partners and bibliography review of included documents. We quantified (existence, frequency) predetermined codes of interest in relation to end-of-life/palliative/care of the dying/bereavement, homecare workforce, and support and training.

Where homecare worker codes were contextually situated, we extracted and tabulated the surrounding text and examined for reference to training and support. We subjected extracted text content to framework analysis through the lens of Bronfenbrenner’s Adapted Ecological Systems Theory.

**Results:** 1,464 homecare worker codes were identified in the 36 included documents, but only 72 times/17 documents in the context of end-of-life care. In the context of end-of-life care and homecare workers, education and training codes were present 3 and 35 times respectively. The need for end-of-life education and training was recognised, but in general, little detail about delivery and implementation was given (e.g., whose responsibility; funding; minimum standard and content).

**Conclusions:** The homecare worker role in end-of-life care is poorly recognised in national policy. Little guidance is provided regarding minimum training standards or delivery. Given an ageing population globally and expected increased demand for end-of-life homecare, national and international policy guidance should include this workforce.

**KEY MESSAGE BOX:** *What was already known?:*

- Social homecare workers (variously known as personal assistants/aides or helps) play a significant role in providing end-of-life care for individuals in their own homes.
- The demand for homecare is increasing due to an ageing population globally and access to good quality provision of end-of-life care is a priority to support community-based care.

*What are the new findings?:*

- There is a policy gap regarding homecare workers in the context of end-of-life care, with little progress since 2010.
- Few policy documents recognise the role of homecare workers in providing end-of-
- life homecare and there is little evidence that previous highly relevant policy recommendations (2010) have been incorporated in more recent current policy or enacted in practice.
- The need for homecare workers education and training in the context of end-of-life care is recognised, but few details about minimum content and standards, or delivery are given.

*What is their significance?:*

- Although this is a UK-based study, given the ageing population internationally with likely increased need for care at the end-of-life, training, role definitions, and support to enable the homecare worker workforce to provide end-of-life care has relevance as a clinical and social care practice priority around the world.
- Policy guidance around end-of-life care should i) include the social homecare workforce; stipulate minimum training requirements in relation to end-of-life care, and ii) integrate this support and training into routine job descriptions, and as part of the working day.
- Improved training and support is an important step to increase the quality of end-of-life care provision.

## INTRODUCTION

Social homecare workers (also known as personal aides/assistants/helps) play a crucial role for individuals wishing to be supported in their own home at the end of their lives and to avoid residential care or hospital admissions where possible (1-3). The size of the adult social care workforce in England (including residential/domiciliary care workers, community and day care workers, and personal assistants) is greater than 1.5 million, increasing yearly (4, 5).

The Homecare Association (UK membership group which supports homecare providers) estimates that 249 million hours of care are delivered annually in England (6). The demand for homecare is increasing as more people are living longer and with multiple chronic conditions; the social care needs of older people are expected to double by 2040 (7). Similarly, assuming population trends continue, the numbers of deaths at home will increase by nearly 100% in this timeframe, requiring a doubling of community-based end-of-life care service provision (7). To address these growing challenges, it is important that access to an appropriately skilled community homecare workforce is prioritized. However, evidence suggests that homecare workers are inadequately trained to deliver end-of-life care and are often isolated and underappreciated (8). This demographic change is being mirrored in many places around the world, with the World Health Organisations urging readiness to ensure their care systems are ready to accommodate this shift (9).

To help address the current challenges facing the social care sector (poor recruitment, retention, pay and conditions), the UK Government plans to professionalise the social care sector which includes developing the knowledge and skills of homecare workers (10) and has recently announced a further major commission to address this growing and critical issue (11).

We aimed to determine current UK health and social care policy priorities and gaps regarding end-of-life care provision by homecare workers, including support and training for the homecare workforce.

## METHODS

We conducted a document review using quantitative and qualitative content analysis to develop a broad picture of current end-of-life carepriorities in social homecare (e.g., if and how it is present in UK health and social care policy) and to identify gaps in policy guidance. This study is part of a wider multi-methods project that aims to improve the quality and sustainability of person-centred end-of-life care provided by homecare workers and involves the use of interviews with homecare clients and carers, homecare workers, community practitioners, homecare managers and commissioners (12).

### Data collection and analysis

We applied Dalglish and colleagues’ systematic approach to document review in health policy research using the READ (Readying, Extracting, Analysing, Distilling) methodology (13) to guide our analysis. The approach consists of four steps: 1) preparing the documents for analysis, 2) extracting relevant data, 3) analysing extracted data, and 4) distilling findings. We analysed extracted text through the lens of Bronfenbrenner’s Adapted Ecological Systems Theory (14): the micro (individual), meso (interactions), exo (services/systems), macro (societal), and chrono (changing needs/complexity over time) context for end-of-life homecare provided by homecare workers; and examined if, and how, it is present in UK health and social care policy.

#### Preparing materials

Policy documents were identified through a scoping of legislation and policy guidance documents by team members, augmented by their networks and our study partners (Bradford Metropolitan District Council, Hull City Council, Skills for Care, Cera Care, London Association of Directors of Adult Social Services). This policy review is part of a larger study including interviews with homecare workers, homecare managers and commissioners (12). As part of the interviews, we also asked participants and stakeholders to suggest key policy documents that they used in their care provision. Lastly, we screened the bibliographies of included documents. Where a document had repeated updates, only the version in current use was included. The last search for documents was completed on 30^th^ June 2024. Titles of included documents were searched online to check for any updated versions on 28^th^ April 2025.

For inclusion in the review, documents had to meet the following criteria: currently in use; relevant to health/social care; direct impact on practice and service delivery or policy and practice development; statutory and non-government organisations.

#### Extracting the data

Documents were read (KM) to assess relevance, authenticity, agenda and biases, audience, tone and style, and purpose (15). Information on the title, author, year of publication (see Table 1), purpose and audience were extracted and tabulated. We then (KM and HE) electronically searched each document, quantifying the existence of predetermined codes of interest relating to end-of-life care, palliative care, support, training, and homecare workers (see Table 2).

**Table 1.**
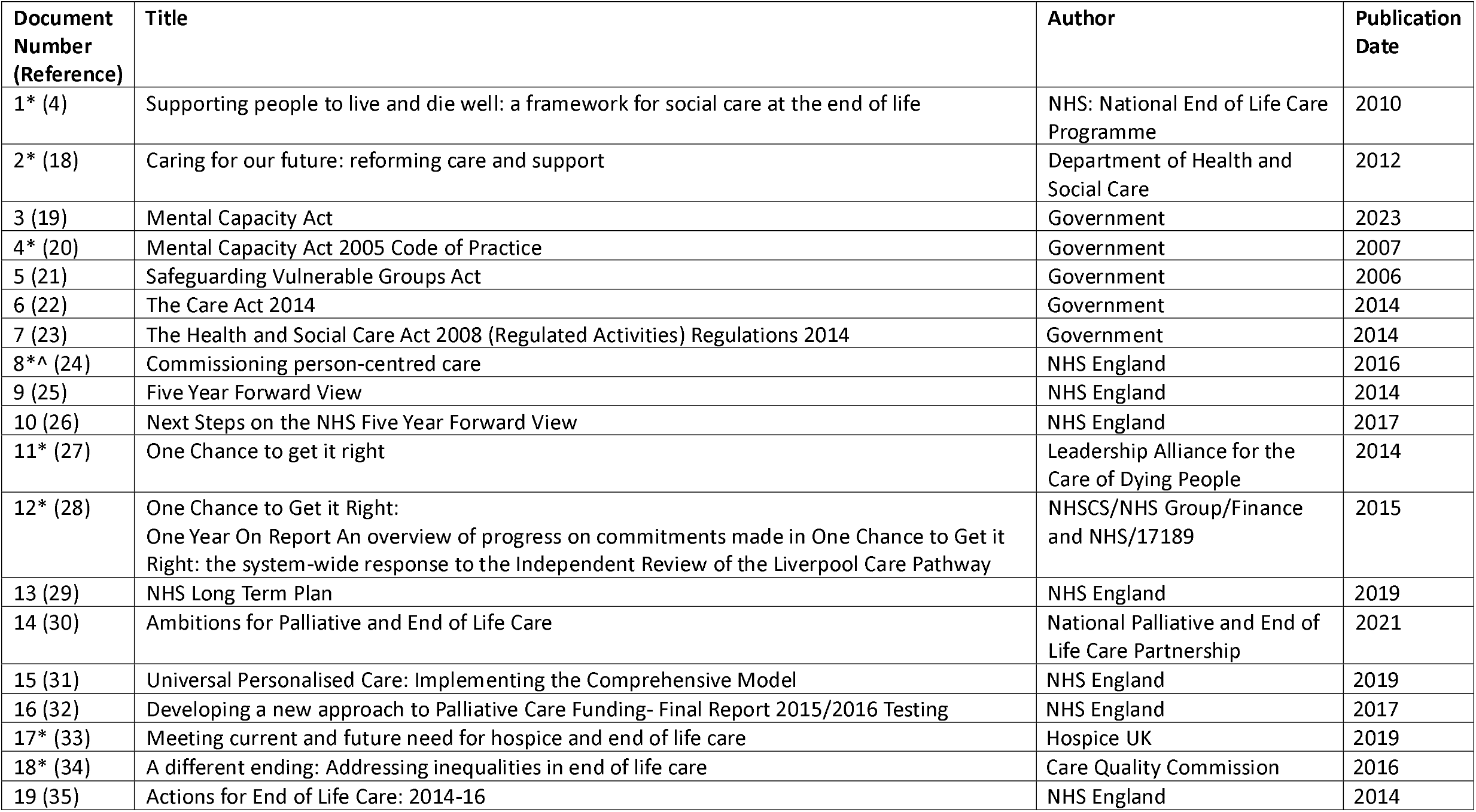
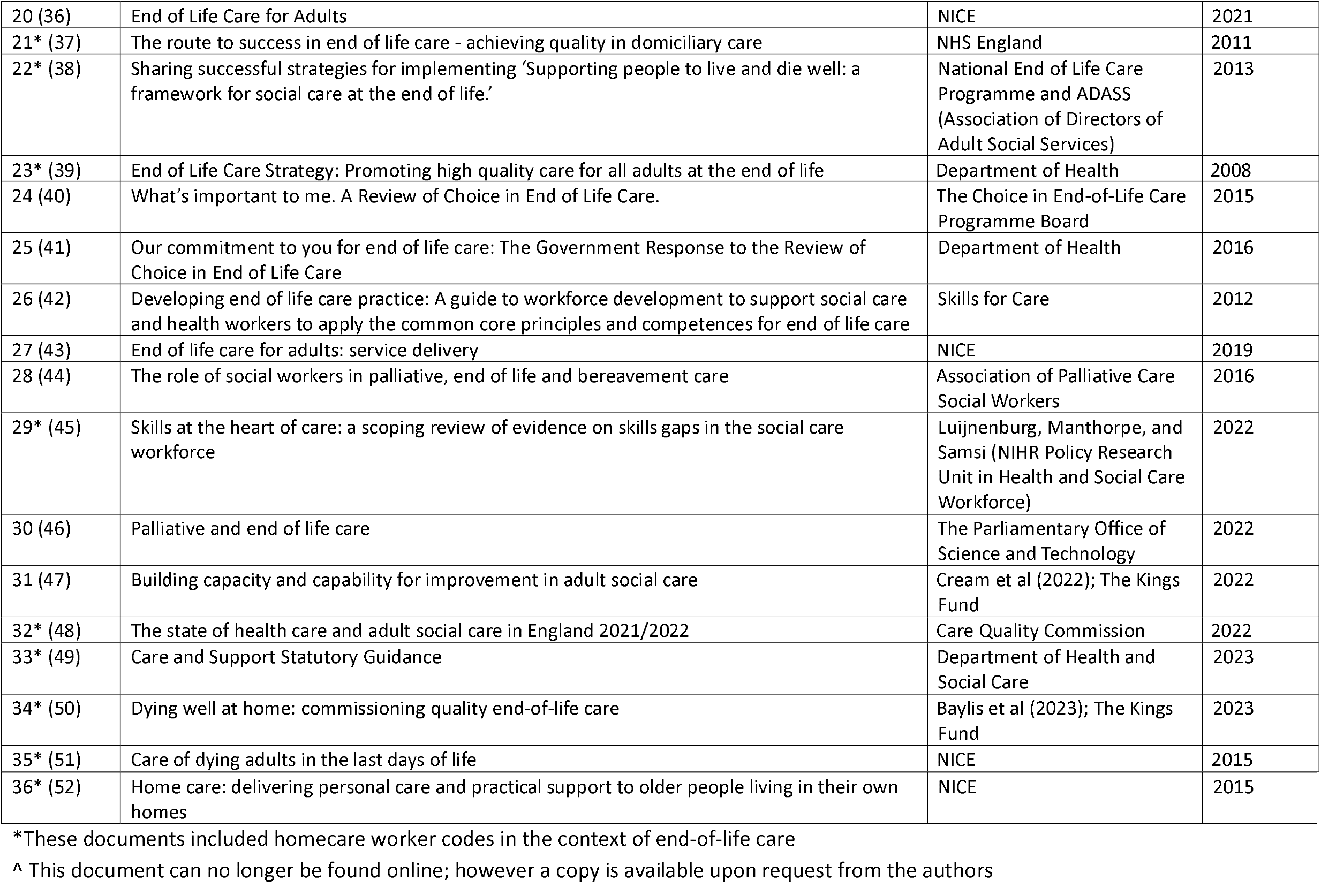
Table of Included Documents.

**Table 2.** Codes of Interest.

| **Key search terms** |  |
| --- | --- |
| End of life care | "End-of-life"/"End of life"/"EoL"* |
| Palliative care | "Palliative"<br>"care of the dying"/"care-of-the-dying"<br>"dying" |
| Bereavement | "bereavement"<br>"grief" |
| Homecare | "Homecare"<br>"Home care"<br>"Domiciliary care" |
| Workforce Knowledge | "education" |
| Workforce Training | "training" |

\*Alternative ending of words accepted

When homecare worker terms were seen in relation to end-of-life care terms, we tabulated the frequency of these codes. Where sentences were repeated verbatim in different parts of the document, codes within repeated sentences were not counted. Surrounding textual contexts were extracted and examined further for reference to training and support. These quotes were tabulated, and duplicates removed.

#### Analysing the data

Frequencies were reported using counts and percentages. Codes per paper were presented as medians, quartiles and ranges. The extracted text (quotes) was combined into one document to determine overall organisational views and priorities, and gaps in workforce support and training relevant to homecare workers providing end-of-life care at home. Quotes were subjected to Framework Analysis (16, 17). They were assigned new codes which were defined and summarised, grouped by theme and mapped onto Bronfenbrenner’s Adapted Ecological Systems Theory theoretical framework (14).

#### Distilling the findings

Findings were interpreted through the lens of Bronfenbrenner’s adapted theoretical framework (14) considering the homecare worker as the centre of the microsystem (the needs and characteristics of the homecare worker). The mesosystem (homecare worker interaction with others, e.g. clients, carers, and other health care professionals), exosystem (service and system level factors), macrosystem (societal factors), and chronosystem (changing needs and complexities over time) were considered in relation to the homecare worker (see Figure 1).

**Figure 1.**
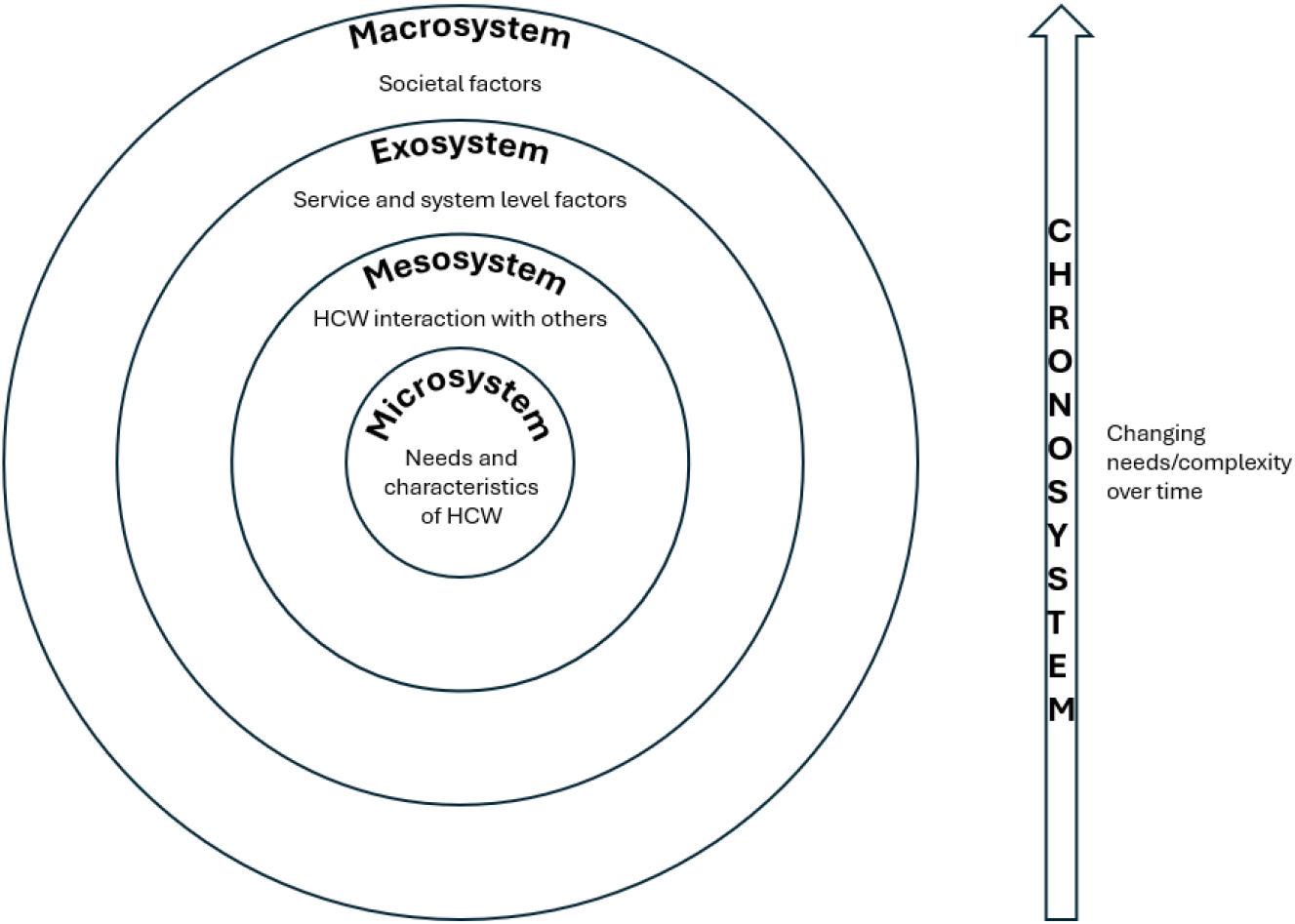
Bronfenbrenner’s adapted theoretical framework applied to homecare workers

## RESULTS

Thirty-six documents were included in the analysis (See Table 1). Codes were quantified across all documents (see Table 2 for Codes of Interest and Table 3 for Frequency of Codes). Homecare worker codes were found 1,464 times across 36 documents (homecare/home care 1392 times in 17/36; domiciliary care 72 times in 14/36).

**Table 3.** Frequency of codes across all papers.

| **Codes of interest (phrases)** | **Number of papers with codes, n (%)** | **Code frequency all papers (n)** | **Codes per paper, median (Q1, Q3) Range** |
| --- | --- | --- | --- |
| End of Life/End-of-life/EoL | 30/36 (83.3) | 4449 | 43.5 (2, 222) 0 – 638 |
| Palliative | 30/36 (83.3) | 1454 | 13 (2, 55.5) 0 – 263 |
| Care of the dying/care-of-the-dying | 11/36 (30.5) | 104 | 0 (0, 1) 0 – 55 |
| Dying | 25/36 (69.4) | 1400 | 9 (0, 53.3) 0 – 506 |
| Bereavement | 21/36 (58.3) | 296 | 2.5 (0, 7) 0 – 92 |
| Grief | 6/36 (16.6) | 22 | 0 (0, 0) 0 – 9 |
| Homecare/Home care | 17/36 (47.2) | 1392 | 0 (0, 5) 0 – 1262 |
| Domiciliary care | 14/36 (38.8) | 72 | 0 (0, 1.75) 0 – 16 |
| Education | 32/36 (88.8) | 810 | 10 (2.25, 23.5) 0 – 131 |
| Training | 33/36 (91.6) | 1523 | 16 (7.25, 53.5) 0 – 231 |

Q1 – quartile 1; Q2 – quartile 2; n - number

The frequency of homecare worker codes in the context of end-of-life care, and training codes in these documents including homecare worker codes are shown in Table 4.

**Table 4.** Codes of interest and frequency, in context of home care workers and end-of-life (n=17)

| **Codes of interest (phrases)** | **Number of papers with codes, n (%)** | **Code frequency all papers, n** | **Codes per paper, median (Q1, Q3) Range** |
| --- | --- | --- | --- |
| End of Life/End-of-life/EoL | 11/17 (64.7) | 43 | 2 (0, 4.5) 0 – 9 |
| Palliative | 4/17 (23.5) | 11 | 0 (0, 0.5) 0 – 6 |
| Care of the dying/care-of-the-dying | 0/17 | 0 | 0 |
| Dying | 5/17 (29.4) | 6 | 0 (0, 1) 0 – 2 |
| Bereavement | 1/17 (5.9) | 2 | 0 (0, 0) 0 – 2 |
| Grief | 0/17 | 0 | 0 |
| Homecare/Home care | 9/17 (52.9) | 48 | 1 (0, 5.5) 0 – 13 |
| Domiciliary care | 8/17 (47.1) | 24 | 0 (0, 2) 0 – 8 |
| Education | 2/17 (11.8) | 3 | 0 (0, 0) 0 – 2 |
| Training | 9/17 (52.9) | 35 | 1 (0, 3) 0 – 8 |

Q1 – quartile 1; Q2 – quartile 2; n - number

### Text analysis

The need for homecare worker education and training in the context of end-of-life care was emphasised in these documents, mainly through identification of training gaps. However, except for one document (Document 1, 2010) (4), there was little clarity regarding homecare worker role boundaries at this time and how to specifically embed end-of-life care training into homecare worker training (e.g. whose responsibility, how funded, minimum standards). Our findings are now described in the context of Bronfenbrenner’s adapted theoretical framework (14), with illustrative quotes (see Supplement Table 1 for details and themes [experiences, support needs, training needs, skills needed, and service provision/development]).

### Microsystem (needs and characteristics of Homecare workers)

The policies that mentioned homecare workers in the context of end-of-life all acknowledged their integral and important role in providing care to patients/clients approaching end-of-life. Homecare workers are a necessary part of the end-of-life care community workforce, but they feel undervalued by society – a persistent issue post-pandemic, unlike care home workers where this has improved somewhat.

> *“The whole social care workforce, including domiciliary and residential care assistants, social workers and other professionals such as occupational therapists and physiotherapists, has a crucial role in supporting individuals, their families and carers at the end of life*.*” Document 1 (4)*

> *“Homecare workers are often the staff who spend most time with people who die at home during their final weeks. However, we heard that they were not well engaged in local approaches to end-of-life care, and that the pandemic had not led to greater understanding and valuing of their role in end-of-life care in the way that it had for care homes*.*” Document 34 (50)*

Documents showed that homecare workers delivering end-of-life care have a range of support needs including having a team member with responsibility for the welfare needs of their homecare workers. Peer support/team working, as well as emotional support, is also needed to support homecare workers in their delivery of end-of-life care and to develop their own skills.

> *“Homecare workers identified peer support and team working as strategies to support their work, and wanted opportunities to develop in their profession, particularly in specialising in dementia care*.*” Document 29 (45)*

However, homecare workers often have inadequate training (e.g. lack of formal qualifications/practical training). The documents evidenced that subject specific training needs are required which cover basic awareness and understanding of end-of-life in domiciliary settings, understanding of role specific models (e.g. Dementia Champions), clinical skills (e.g. continence management, mouth and dental care, palliative care skills), knowledge and understanding of specific conditions, awareness of dignity and respect, observational skills, and interpersonal/communication skills.

> *“Communication skills training aimed at end-of-life conversations is therefore particularly important for all levels of social care worker, including care assistants in domiciliary and care home environments, to develop confidence and refine the skills required to discuss end of life care needs with individuals and their families*.*” Document 1 (4)*

> *“There is also a need to acknowledge and develop the observational skills of spotting ‘soft signs’ of deterioration among homecare users…As yet, there is little evidence around the use of soft signs in homecare, but in care homes such skills are emerging as important to foster*.*” Document 29 (45)*

Minimum quality and safety standards for home-based care in general are available, and although they are not explicitly linked to end-of-life care, it was recognised that all are relevant to the provision of good end-of-life care (37). However, documents indicated that homecare workers also need to develop end-of-life care-specific skills such as understanding holistic approaches to advance care planning, understanding of ed-of-life in the domiciliary setting, dignity and respect, how to discuss end-of-life care preferences, and contribution to the wider care team’s planning; the homecare worker is an important part of this aspect of care.

> *“Social care has an important role to play in linking the holistic assessment of need to a holistic approach to advance care planning. This may include facilitating conversations with other professionals at a time that is right for the individual service user, which may be considerably earlier than envisaged*.*” Document 1 (4)*

> *“It is vital that care workers can recognise the dying phase and take appropriate action. How someone dies remains a lasting memory for the relatives, friends and care workers*.*” Document 21 (37)*

Delivery of care apprenticeships, minimum training standards, appropriate end-of-life education programmes, and induction programmes for homecare workers were all suggested as ways to address training needs. Robust access to training, along with addressing the current recruitment crisis was seen as a potential solution.

> *“Inadequate training and skills base. Only 7% of domiciliary care workers and 5% of care home assistants hold an NVQ level three qualification that includes (optional) training in support for people at the end of life*.*” Document 1 (4)*

### Mesosystem (interactions between clients, carers and practitioners [Homecare workers and other health and social care professionals])

The social care workforce includes homecare workers provided from care organisations, private and independent care providers, and local authorities. The documents reviewed highlight that it is important for homecare workers, other professionals, and health care professionals across the workforce to work together to ensure high quality delivery of end-of-life care. However, despite homecare workers having vital information for local service planners and providers, they are usually omitted from stakeholder engagement activities and work in relative isolation from the healthcare team. It was also noted that it can be hard to define the homecare workers’ role in relation to other social and healthcare providers, aggravated by poor communication between the social and health care professionals providing the care.

> *“We heard that they were not well engaged in local approaches to end-of-life care…Feeding in information from homecare services is essential if there is to be a whole pathway, end-to-end view of the quality of end-of-life care at home. And focusing on the way these teams work with the health care side of the multidisciplinary team that supports people at the end of life is a key part of developing end-to-end quality improvement plans*.*” Document 34 (50)*

> *“Allowing an individual to die with dignity in the comfort of their own home with their own family around them is a key measure of good end of life care provision. A key challenge for domiciliary home care workers is the extent to which it is appropriate for them to become involved in an individual’s discussions about their personal wishes and preferences for care*.*” Document 21 (37)*

Support for homecare workers needs to be provided by their senior colleagues and managers, which considers their welfare needs. This can include good supervision practices, regular access to informal or ad hoc support and varied training opportunities. However, homecare workers often carry out lone working, limiting opportunities for both support and learning from others in the immediate and extended team. There is currently a lack of documentation or guidance evidencing the need for this support for homecare workers.

### Exosystem (service and system level factors)

Many documents reviewed suggested that role clarification of different health and care workers would be an important first step to providing an integrated service at the regional and national levels. This would provide a common understanding of roles and an appreciation of the work which each carry out.

> *“The NEoLCP [National End of Life Care Programme] should seek to engage with social care umbrella organisations and professional associations … to clarify and articulate the role of residential and domiciliary care workers, housing and other support workers, occupational therapists (OTs), physiotherapists and nurses in integrated care teams, for example learning disability nurses*.*” Document 1 (4)*

> *Feeding in information from homecare services is essential if there is to be a whole pathway, end-to-end view of the quality of end-of-life care at home. And focusing on the way these teams work with the health care side of the multidisciplinary team that supports people at the end of life is a key part of developing end-to-end quality improvement plans*.*” Document 34 (50)*

Additionally, it was recognised that for homecare workers to function within their organisations and be retained, they need adequate pay (at least minimum wage), to be employed by care providers with appropriate skill sets within their organisations, better support structures, and financial stability. One solution, at both the levels of the exosystem and mesosystem, is for services employing the homecare workers to have greater funding for training and development activities within worktime.

> *“The Low Pay Commission’s 2012 report highlighted a possible relationship between 15-minute home care appointments and some care workers being paid below the national minimum wage. It is the responsibility of employers to ensure that staff are paid at least the national minimum wage, and the Government will work with the Low Pay Commission and local authorities to understand and challenge the reasons behind cases of non-payment of the minimum wage by employers to care workers*.*” Document 2 (18)*

> *“Few care staff possess specialist palliative care skills while some care agencies will not be able to manage people with physical conditions or those approaching end of life, due to funding issues and/or skill sets within that organisation*.*” Document 21 (37)*

Some policies highlighted the need for greater understanding of the complexities of end-of-life care and the professionals involved (amongst other social service practitioners/organisations), so they are aware of wider care network factors that may impact their delivery of care.

> *“[regional council] has funded the pilot of an 18-month social care project run by [name] Hospice, to deliver domiciliary care to people in their last year of life. This collaborative project is raising awareness of end-of-life care issues amongst social service staff who liaise with the project*.*” Document 22 (38)*

We found several possible solutions to meeting the training needs of homecare workers which can be provided within the wider context of the exosystem (from links with other agencies, sources outside their own) including: engagement with further and higher education providers, using Domiciliary Care Frameworks to determine appropriate and high quality end-of-life care, meeting essential/minimum care standards, using role specific models (e.g. Dementia Champions), embedding and accrediting skills within training programmes, utilising care certificates, and agencies such as Skills for Care (53) (the strategic workforce development and planning body for adult social care in England).

Specific end-of-life care training needs are recognised (see Microsystem section above); documents show that care providers will need to demonstrate to national quality regulators (e.g., Care Quality Commission, England) how they ensure their staff are competent in “*assessing and responding to the holistic and changing needs of individual dying people and their families*.” *Document* 11 (27), as part of person-centred care provision at end-of-life.

Solutions to meeting training needs can be provided within the wider context of the mesosystem, from other organisations (e.g. Skills for Care) and professionals providing End-of-life Care for All programme online training (e-ELCA, a UK-based e-learning library providing training and education resources for the health and social care workforce), care apprenticeships, and care certificates. Other training in dignity and respect, observational skills, decision making skills, and team working would allow minimum standards of training to be met. Appropriate end-of-life care education and induction programmes for homecare workers could be delivered. Where possible, skills assessment could be embedded within training programmes and accredited allowing homecare workers to gain the recognition they deserve. It was suggested that training could also be provided by other professionals and care organisations within the wider care sector.

> *“Local authorities should consider, in particular, how to encourage training and development for the care and support workforce, including for the management of care services, through, for example, national standards recommended by Skills for Care*.*” Document 33 (49)*

> *“There should be engagement with further and higher education providers and the diverse range of independent training organisations to influence the development of curricula and programmes so as to provide a range of opportunities at all levels for social care workers to access education and training in end-of-life care. This will extend to independent providers of domiciliary care workers or care assistants and carers who work within care home settings as well as the generalised social care workforce. It should include engaging with the Social Work Reform Board and the redevelopment of social worker training at qualifying, post-qualifying and advanced levels; Skills for Care could include a priority target in allocating their Training Strategy Implementation funding*.*” Document 1 (4)*

It was clear from the review that formal and mandatory training and support is needed for all the social care workforce. It was suggested that this could be provided by local authorities or other agencies and could include national standards. Consistency of training would enable homecare workers across the workforce to have reliable training and support to deliver high quality end-of-life care. Recommendations were made that employers should ensure that homecare workers receive appropriate training (through induction or other continuing professional development programmes), and appropriate funding must be available to support such training. Financial pressures on services need to be considered (such as issues relating to pay/conditions, and travel costs for homecare workers).

> *“When social care is commissioned by, but not directly provided by local authorities, how is consistency in training managed so all care agency staff are equipped to deliver good end of life care?” Document 8 (24)*

> *“Local authorities should consider encouraging the training and development of care worker staff to at least the standard of the emerging Care Certificate currently being developed by Health Education England, Skills for Care and Skills for Health*.*” Document 33 (49)*

> *“…and have regard to funding available through grants to support the training of care workers in the independent sector*.*” Document 33 (49)*

In relation to service provision and development, the review recognised that the type of support needed should be able to meet all, new, and changing needs of home care services.

> *“There are also questions about whether community-based services that are already under strain are equipped to support a growing number of people, often with highly complex needs, to die at home*.*” Document 34 (50)*

### Macrosystem (societal factors)

The review highlighted that among the public, there should be a greater awareness and understanding of domiciliary care services and the role they play in the provision of end-of-life care. As noted above, society does not seem to value homecare workers for the crucial role they play in end-of-life care although the COVID-19 pandemic improved this perception for care home staff.

> *“The general public is often unaware of the role that social care can play in improving people’s experiences of dying, for example through access to domiciliary care services, acting as an intermediary with other services, and facilitating the making and upholding of individual choices*.*” Document 1 (4)*

To support homecare workers, appropriate action is needed to overcome challenges in the social care system, so appropriate end-of-life care can be delivered to individuals and families. In relation to service provision and development, the review highlighted that financial pressures on services need to be considered (such as issues relating to pay/conditions, and travel costs for homecare workers). This would alleviate poor working conditions relating to these factors and enable organisations to better meet the end-of-life care needs within their communities by recruiting and retaining more staff, but this may depend on the extent to which society values their role relative to other financial pressures in the public sector.

> *“To date, a range of challenges and barriers has tended to limit the contribution of social care. Action is now needed to overcome these challenges and barriers to ensure social care fulfils its potential to enhance the support provided for individuals and their families at the end of life*.*” Document 1 (4)*

> *“In terms of recruitment, as well as the sheer lack of applications, both care home and homecare providers reported challenges including candidates lacking necessary skills and experience and issues with pay and conditions*.*” Document 32 (48)*

> *“Petrol and diesel prices have also had an impact on homecare staff who rely on a car to get them to their visits. Our adult social care workforce survey showed that, of the homecare services that provided information about retention challenges, nearly a quarter (23%) reported challenges related to the increased cost of petrol*.*” Document 32 (48)*

### Chronosystem (changing needs and complexities over time)

Documents suggest that there are insufficient resources to support social care packages required for adequate care. This is further compounded by staff/workforce shortages. This can lead to care packages being handed back to local authorities as they cannot be delivered with limited available hours of homecare. There can also be delays in discharging people from hospital due to a lack of homecare support. For people at the end of life, the timeframe to discharge someone to be able to die at home may be short, and deaths in hospital can occur if social homecare provision is not sufficiently resourced to respond quickly. Workforce shortages arise from recruitment and retention difficulties, lack of homecare worker skills and experience, and issues with pay and poor working conditions.

> *“There are large numbers of patients who are stuck in hospital longer than they need to be, due to a lack of available social care packages*…*This is due, to a large extent, to severe staff shortages in adult social care*.*” Document 32 (48)*

Evidence suggests that given the increase in older adults with increasingly complex multiple long-term conditions requiring homecare at end-of-life, to enable high quality provision of services, a better service configuration is needed to provide longer and more sustainable appointment times. Metrics (for example, collecting and acting on user feedback, and documenting length and frequency of visits) should inform quality of services, and allow for continuous improvement. Local authorities and other organisations providing care should consider capacity, continuity, and flexibility of services to support wellbeing and fundamentals of care for those delivering and receiving end-of-life care, recognising that people with non-cancer conditions will also require end-of-life care; many end-of-life care services still only take cancer into account when estimating workforce requirements.

> *“End of life care was not discussed, because it wasn’t cancer. I have cared for people with end stages of COPD and renal failure – neither got any end-of-life care and their lives and mine as carer were adversely affected, even though it was known they would shortly die. Only cancer sufferers appear to be included in end-of-life care*.*” Document 18 (34)*

## DISCUSSION

### Main findings/results of the study

The policy documents reviewed recognise that homecare workers play an integral role in the provision of end-of-life care for those dying at home. However, this is explicitly discussed in relatively few policy documents in current use. In general, where the included policy documents did mention homecare workers in the context of end-of-life care, the topic was superficially covered. Unmet subject specific training needs were acknowledged, along with a lack of formal skills and qualifications. Solutions suggested in the documents reviewed included formalised training, shared learning with other organisations, embedding domiciliary care frameworks and essential standards into training programmes, and accrediting training so homecare workers feel rewarded in their role (this could also ensure validity and quality of training). In addition, adequate pay and sustainable funding for training are required for homecare worker organisations. Greater public awareness is needed about homecare workers and their roles, as they are often overlooked in their provision of end-of-life care; this may improve/support change and wider societal appreciation of these roles. Any homecare worker service must be flexible enough to recognise rapidly changing needs of people with deteriorating disease and respond to urgent care needs, such as hospital discharge to enable death at home.

One document (Document 1) (4) did provide detailed context regarding homecare worker role boundaries and suggested recommendations regarding how to specifically embed end-of-life care training into homecare worker training (including whose responsibility it is, issues related to funding, the minimum standard). There was content regarding gaps in training and the support that homecare workers require, along with issues relating to service provision of end-of-life care. Although published in 2010, there was little evidence that these strong recommendations had been implemented in practice or incorporated into more recent documents.

### What this study adds

Our findings are consistent with the broader evidence concerning homecare workers working in end-of-life care. A recent rapid review (8) highlighted that homecare workers have extensive training and support needs, require emotional and educational support in managing complex situations encountered within end-of-life care, tend to work in isolation, and are often overlooked and undervalued by other social and healthcare workers, and society more broadly. Our interview data from the SUPPORTED study (12) (paper in preparation) indicates that the availability of this is patchy with some receiving minimal or no end-of-life care training and required to use their own initiatives and resources to fill these gaps. Further, a recent mixed-methods systematic literature review (54) of homecare workers’ training and psychosocial needs when supporting people living with dementia, also highlighted four main themes with which our findings resonate: unmet training and education needs; social isolation; emotional attachments and distress; and working with families. These unmet needs were shown to have a negative impact and require condition-specific training (i.e. Dementia), emotional/peer support, and support to manage relationships with clients and families. The ongoing challenges for homecare workers described in these recent reviews, are the same concerns described in detail in the 2010 policy document. This indicates a policy-practice gap, which, given a deepening workforce crisis and a growing demand from an ageing population, is widening (11, 55). Service provision gaps relate to the lack of available social care packages and workforce shortages, which could be resolved by better configuration of services, providing more sustainable and consistent care (55, 56). Further, there is a growing literature which alongside our own findings (12) (paper in preparation) demonstrates the emotional impact of bereavement experienced by homecare workers providing end-of-life care (57, 58). In contrast, the policies reviewed here heavily neglected this important aspect of homecare worker support, which can be anticipated to alleviate the emotional impact of working with people close to and at the time of death. This support may contribute to retention of homecare workers, over and beyond any contributions made by improved training and remuneration.

Staffing and retaining an adequate workforce, low appeal of care work, poor pay rates and competition from other companies, rising demand and reduced funding for councils, and contracts being handed back to councils (due to the uncertainty of future homecare and its fragile market) (6), are all factors impacting on the provision of good quality end-of-life care by homecare workers and care agencies. Our included policy documents reflected these issues throughout.

Current policy and guidance for homecare workers is lacking in that there are some documents which provide good advice and guidance (Document 1, 2010) (4), but these are out of date and health and social care structures have changed in this time. The recent Nuffield Trust report (56) has reflected on the failing UK social care system and a lack of prioritisation within Government (partly due to the Covid-19 pandemic) and has called for a comprehensive reform of the entire system. Of note was the temporary shift in perceptions of homecare workers (and similar roles) to one of value and positivity during the Covid-19 pandemic, demonstrating that the status of homecare workers fluctuates depending on wider societal issues. This exemplifies a need for better training which can validate the complexities of the role.

Further, a recent paper discussing adult social care reform referred to the years from 2010 as a ‘lost decade’ for adult social care; albeit not specific to end-of-life care. Unmet need, self-funding, poor quality care, and greater pressures on staff and families were cited as a call for urgent action (55). Changing health and social care structures would provide opportunity and means to create better end-of-life care provision for those dying at home.

The independent commission into adult social care (11), announced by the UK Government in January 2025, recognises the crisis in the social care workforce and the need for significant reform and new models of care. The core tasks of the commission are centred around: i) the best use of technologies in enabling integrated care (including joined up digital systems), effective communication between service providers, care professionals and families, thus supporting people to live (and stay) at home independently; and ii) professionalisation of the social care workforce including training to take on delegated healthcare activities, expanding the national career structure, and associated recognition and reward. This commission will have the potential to recognise and close the glaring gap identified in this policy review on social home care delivery at the end-of-life, thus addressing the ongoing silence in these contexts.

### Strengths/weaknesses

To identify the organisational views and priorities reflected in national policies related to homecare workers providing end-of-life care at home, we conducted a document analysis. This is a common and useful systematic method for reviewing and/or evaluating documents and has various uses, including to provide context, generate questions, and triangulate various types of data (15).

Further, the application of Bronfenbrenner’s Adapted Ecological Systems Theory (14) helped organise our data, strengthened the interpretation of findings, and helped contextualise our understanding of the current landscape and challenges in supporting homecare workers to provide good quality end-of-life care.

Our search for policy documents was guided by team members, their networks, stakeholders, and study partners. It is possible that other documents were missed, or published as this work was being carried out/written up, but as we included those from key national drivers and sources, we consider we are unlikely to have missed any central documents.

Terminology for codes was based on those used within a UK policy context (e.g., home care worker), and therefore other terms may be more applicable within a wider, international context.

### Implications for policy and practice

Although UK-specific, we believe many of our findings have relevance and are applicable irrespective of health and social care delivery model in other countries facing similar demographic change. Our rapid review (8) exploring the needs and experiences of homecare workers included papers from Japan, the United States of America, Canada and Sweden as well as the UK, and found similar issues relating to inadequately training, isolated work patterns and poor levels of appreciation by others.

- Homecare workers should be trained appropriately in the delivery of end-of-life care at home (skills assessment could be embedded within training programmes with appropriate accreditation, allowing homecare workers to gain recognition for their work).
- Homecare workers should be included in relevant policy and service development.
- Homecare workers should be included in interprofessional collaboration and working within community-based end-of-life care practice.
- The health and social care sector must consider questions about providing sustainable, flexible and affordable services, depending on social and healthcare service models.
- We need to consider the best metrics to use as markers of quality care.
- Societal attitudes – we need to consider what value is placed on adequate end-of-life care at home, and those that provide it.
- Recognition that robust homecare end-of-life care services are integral and need to be well integrated within the broader end-of-life care system, and therefore have a significant impact on all social and healthcare services.
  - e.g., inappropriate use of hospital beds (the accompanying healthcare crisis cannot be resolved without addressing social homecare (11, 59)).

### Implications for research

This study is part of a wider multi-method project that aims to improve the quality and sustainability of person-centred end-of-life care by homecare workers, identify training and support needs, and develop training resources, and involves the use of interviews with homecare clients and carers, homecare workers, community practitioners, homecare managers and commissioners (12). Future work should include policy review from other countries to look for good examples, and possible solutions.

## CONCLUSION

Within UK current national policy guidelines, we identified priorities and gaps regarding end-of-life care provision by homecare workers. These relate heavily to support and training for the homecare workforce. There is a lack of standardised minimum training requirements for homecare workers. To increase the quality of end-of-life care provision for those dying at home, training should be provided in a formalised and accredited manner to homecare workers. Homecare workers would also benefit from increased support from, and integrated working with, peers, colleagues, and other health and social care professionals from across the workforce.

The health and social care workforce needs better configuration of services including greater communication, more joined up working between providers and professionals, and financial sustainability, to allow homecare workers to deliver appropriate end-of-life care. Further, as homecare workers are an integral source of end-of-life care provision for individuals dying in their own home, they should be included in policy development and service delivery decision making.

The role of homecare workers in providing end-of-life care is poorly recognised in UK policy. Where it is acknowledged, little detailed guidance is provided regarding minimum training standards or delivery models. Given similar problems internationally due to an ageing population, and that many people wish to receive end-of-life care at home, policy guidance globally should include this crucial workforce.

The ongoing Government health policy push for services to move from hospital to community care offers an opportunity to recognise and address the critical need for effective delivery, and support for, end-of-life care at home.

## CONFLICTS OF INTEREST

The authors have no conflicts of interest to declare.

## ACKNOWLEDGEMENTS

The authors want to thank Ms Kathryn Harvey, administrator for the SUPPORTED study, who helped with data extraction for this report, and with overall administration of the project.

## DATA AVAILABILITY

The data used for this review are all in the public domain. See Table 1 for details of the documents used.

## FUNDING

The author(s) disclosed receipt of the following financial support for the research, authorship, and/or publication of this article: This study was funded by the NIHR HSDR Programme (Ref: NIHR135128) as part of the SUPPORTED project, a wider multi-method project that aims to improve the quality and sustainability of person-centred end-of-life care by homecare workers.

The views expressed are those of the author(s) and not necessarily those of the NIHR or the Department of Health and Social Care.

## ETHICS STATEMENT

Ethics approval was not required for this study as all data was in the public domain.

## Notes

### Competing Interest Statement

The authors have declared no competing interest.

### Funding Statement

This study/project is funded by the NIHR HSDR Programme (project reference NIHR135128). The views expressed are those of the author(s) and not necessarily those of the NIHR or the Department of Health and Social Care'

