## Supplemental Table for "Policy gaps regarding social homecare in the context of end-of-life; a policy document analysis"

**Supplementary Table 1. SUPPORTED Policy Document Review**

| **Theme** | **Microsystem (needs and characteristics of HCWs)** | **Mesosystem (interactions between clients, carers and practitioners [HCWs and other health and social care professionals])** | **Exosystem (service and system level factors)** | **Macrosystem (societal factors)** | **Chronosystem (changing needs and complexity over time)** |
| --- | --- | --- | --- | --- | --- |
| **Definitions** | - Care workers   *HCWs provide immediate EoLC to patients/carers.* | - Patients/clients - LA - NHS - Other care providers   *The social care workforce includes HCWs care organisations, private and independent care providers, and local authorities.* | - Domiciliary care - Size of adult social care workforce - Service description; role definition   *Definitions and role clarifications regarding domiciliary care would be advantageous.* | - Public awareness   *There needs to be greater public awareness and understanding of domiciliary care services.* |  |
| **Experiences of HCW’s** | - HCWs play an integral/important role in providing care   *HCWs are a necessary part of the end-of-life care community.* | - Working together (across HCPs) - Working together   *HCW’s work with other providers and practitioners across the workforce.* | - HCWs play an integral/important role in providing care   *HCWs are a necessary part of the end-of-life care community.* |  |  |
| **Support needs of HCW’s** | - Support for HCWs delivering EoL care - Peer support/team working - Emotional support   *HCWs have support needs which help them deliver appropriate EoLC; these needs impact on their ability to deliver care.* | - Solution   - importance of supervision/welfare of others   *HCW’s need good supervision, which considers their welfare needs.* | - Adequate pay (at least minimum wage) - Care providers   - Financial issues   - Skill set - Awareness of EoL care issues amongst social service staff - Peer support/team working - Emotional support - Solution   - Funding for training   *HCW’s need appropriate pay and skills sets, which could be provided by better funding for training.* | - Action needed to overcome challenges/barriers of support at EoL   *Action is needed to overcome challenges/barriers in the social care system, to deliver appropriate EoLC to individuals and families.* | - Solution   - HCWs should be included in policy and service development |
| **Training needs of HCW’s** | - Subject specific   - communication skills   - dignity and respect   - care apprenticeships   - minimum training standards   - appropriate EoL education programmes   - understanding of EoL in domiciliary setting   - induction programmes for care workers   - observational skills   - skills should be embedded and accredited   - continence management   - mouth and dental care   - basic awareness and understanding of end of life in domiciliary setting   - palliative care   - specific conditions   - role specific models, e.g. Dementia Champion - Inadequate training - Minimum training standards   *There are subject specific training needs which should be implemented and upheld for the HCW.* | - Subject specific   - communication skills   - holistic approach to advanced care planning   - minimum training standards   - appropriate EoL education programmes   - induction programmes for care workers   - skills should be embedded and accredited - Adequate supervision - Solution   - e-ELCA online training   - care apprenticeships   - dignity and respect   - meeting standards   - Care Certificate   - Skills for Care   - observational skills   - decision making skills   - team working   - skills should be embedded and accredited   - minimum standards   *There are subject specific training needs (and solutions) which should be implemented and upheld for the HCW.* | - Subject specific   - holistic approach to advanced care planning   - minimum training standards   - skills should be embedded and accredited   - basic awareness and understanding of end of life in domiciliary setting   - role specific models, e.g. Dementia Champion - Solution   - e-ELCA online training   - care apprenticeships   - dignity and respect   - engagement with FE/HE providers   - high quality care   - essential standards   - Domiciliary Care Framework and delivering EoL care   - Role specific models, e.g. Dementia Champion   - Care Certificate   - Skills for Care   - Minimum standards   - skills should be embedded and accredited - Training and support needed for all of social care workforce - Consistency of training - Requires input at local authority level - Employer to ensure appropriate training to health care workers - Funding may be required   *There are subject specific training needs (and solutions) which should be implemented and upheld for the HCW, this can be provided by other professionals across the care sector.* |  |  |
| **Skills needed** | - Holistic approach to advanced care planning - Dignity and respect - How to have discussions about end-of-life care preferences - Understanding of end of life in domiciliary setting   *HCW’s need to develop EoLC-specific skills.* | - Holistic approach to advanced care planning - Dignity and respect - How to have discussions about end-of-life care preferences - Understanding of end of life in domiciliary setting   *HCW’s need to develop EoLC-specific skills.* | - Holistic approach to advanced care planning - Dignity and respect - How to have discussions about end-of-life care preferences - Understanding of end of life in domiciliary setting   *HCW’s need to develop EoLC-specific skills.* |  |  |
| **Service provision/**  **development** |  |  | - Type of support needed - Financial pressures on services   *There are changing needs across the sector which should be considered to support home care services.* | - Financial pressures on services   *Financial pressure on services (e.g. pay/conditions/travel) need to be considered.* | - Lack of available social care work packages/workforce shortages - Quality of services/service provision - Service development – HCWs should be included in policy and service development   *There is a lack of social work packages due to workforce shortages, which impacts the quality-of-service provision of EoLC. HCWs must be included in policy and service development (considering their fundamental role in the provision of EoLC), making them feel more included in decision making.* |
| ***Narrative Summary*** | *Microsystem (needs and characteristics of HCWs)* | *Mesosystem (the interactions between clients, carers and practitioners [HCWs and other health and social care professionals])* | *Exosystem (service and system level factors)* | *Macrosystem (societal factors)* | *Chronosystem (changing needs and complexity over time)* |
|  | Social homecare workers (HCWs) play an integral and important role in providing care to patients/clients approaching end-of-life. HCWs are a necessary part of the end-of-life care (EoLC) community workforce.  There are support needs for HCWs delivering EoLC and these include: having someone on the team whose responsibility it is to consider the welfare needs of their HCWs; peer support/team working from peers/colleagues/other professionals to support HCWs delivery of EoLC and to develop their skills; and emotional support to manage their delivery of EoLC.  HCWs often have inadequate training (lack of formal qualifications/necessary skills), and therefore minimum training standards should be implemented and upheld. Subject specific training needs are required which cover the following: communication skills, dignity and respect, basic awareness and understanding of EoL in domiciliary setting, observational skills, continence management, mouth and dental care, palliative care skills, understanding of specific conditions, and understanding of role specific models (e.g. Dementia Champions).  These training needs can be delivered through care apprenticeships, minimum training standards, appropriate EoL education programmes, and induction programmes for HCWs. Skills assessment could be embedded within training programmes and accredited for HCWs to gain recognition for their work.  Further, HCWs also need to develop skills and understanding of holistic approaches to advanced care planning, understanding of EoL in the domiciliary setting, dignity and respect, and how to have discussion about EoLC preferences. | The social care workforce is imperative in supporting individuals and their families at the EoL. This includes home care workers provided from local authorities, care organisations, and other care providers. It also includes other health and social care professionals (HCPs) within this setting.  It is important for HCWs, other professionals, and HCPs across the workforce to work together to ensure high quality delivery of EoLC.  For this to happen there needs to be good supervision for HCWs provided by their superiors, which considers their welfare needs.  Subject specific training needs are required for HCWs and this includes an understanding of holistic approaches to advanced care planning, which includes working with other professionals within social care to benefit the individual receiving EoLC.  Solutions to training needs can be provided within the wider context of the mesosystem, from other organisations (e.g. Skills for Care) and professionals providing e-ELCA online training, care apprenticeships, and care certificates. Other training in dignity and respect, observational skills, decision making skills, and team working will allow minimum standards of training to be met. Where possible skills assessment could be embedded within training programmes and accredited for HCWs to gain the recognition they deserve.  HCWs also need to develop skills and understanding of holistic approaches to advanced care planning, understanding of EoL in the domiciliary setting, dignity and respect, and how to have discussion about EoLC preferences. These can be provided by other professionals and care organisations within the wider care sector. | Domiciliary care is that which is provided to individuals/families in their own home by a HCW. The size of the total adult social care workforce (which includes residential/domiciliary care workers, community and day care workers, and personal assistants) is greater than 1.5 million, increasing yearly. Clarification for the definitions of different care workers would be advantageous.  HCWs are an important and integral part of the wider system, providing care to individuals and their families at the end of life.  For HCWs to function within their organisations, they need support which includes receiving adequate pay (at least minimum wage) and be employed by care providers who have appropriate skill sets and financial stability. One solution for this would be for the services employing the HCWs to have greater funding for training/services.  HCWs need to have an awareness of EoLC issues amongst other social service staff/organisations in the wider system (outside of their own organisation), so they are aware of other societal factors which may impact their delivery of care.  Support needs extend to peer support/team working and emotional support which can all be provided by HCWs immediate teams or those they are employed by.  Solutions to training needs can be provided within the wider context of the exosystem (from links with other agencies, sources outside their own) including: engagement with further and higher education providers, using Domiciliary Care Frameworks to determine appropriate and high quality EoLC, meeting essential/minimum care standards, using role specific models (e.g. Dementia Champions), embedding and accrediting skills within training programmes, utilising care certificates, and agencies such as Skills for Care.  Formal and mandatory training and support is needed for all the social care workforce; this could be provided by local authorities or other agencies and could include national standards. Consistency of training would enable HCWs across the workforce to have reliable training and support to deliver high quality EoLC. Employers should ensure that HCWs receive appropriate training (through induction or other training programmes).  Local authorities should consider encouraging training/development for the care workforce.  Appropriate funding must be available to support the training of HCWs.  HCWs need to develop skills and understanding of holistic approaches to advanced care planning, understanding of EoL in the domiciliary setting, dignity and respect, and how to have discussion about EoLC preferences. These can be provided by other professionals and care organisations within the wider care sector.  In relation to service development, the type of support needed should be able to meet all, new, and changing needs of home care services. This should consider whether the service is equipped to deal with the complex needs of EoLC.  Financial pressures on services also need to be considered (such as issues relating to pay/conditions, and travel costs for HCWs).  Policy and guidance to services for HCWs is lacking in that there are some documents which provide good advice and guidance (2010), but these are old, have not been updated, and NHS/social care structures have changed in this time. | Amongst the public, there should be a greater awareness and understanding of domiciliary care services, and the role they play in the provision of EoLC.  To support HCWs, appropriate action is needed to overcome challenges/barriers in the social care system, so appropriate EoLC can be delivered to individuals and families.  In relation to service provision/development, financial pressures on services need to be considered (such as issues relating to pay/conditions, and travel costs for HCWs). This would alleviate poor working conditions relating to these factors and enable HCWs to provide adequate EoLC. | In relation to service provision/  development, there are often a lack of available social care packages resulting from staff/workforce shortages, which results in care packages being handed back to local authorities as they can’t be fulfilled, or people are not able to leave hospital due to a lack of homecare support. Workforce shortages also include challenges to recruit, lack of skills/experience, and issues with pay/conditions.  For high quality provision of services, better configuration of services is needed to provide longer and more sustainable appointment times, minimum wage, better/more consistent training, and good quality of care for all (irrespective of location and disease). Data collection should inform on quality of services and allow for continuous improvement. Local authorities should consider capacity, continuity, and flexibility of services to support wellbeing and fundamentals of care for those delivering and receiving EoLC.  It is important to recognise that HCWs are often overlooked and should be included in policy and service development (considering their fundamental role in the provision of EoLC). This could also be a solution to help with the support needs of HCWs as it may help them feel more included in decision making about EoLC within the social care setting. |

HCWs – home care workers; NHS – National Health Service; EoLC – end of life care; e-ELCA – End of Life Care for All, e-learning for Healthcare;
